# Sensor-free motion registration and automated movement evaluation: Leveraging machine learning for clinical gait analysis in ataxia disorders

**DOI:** 10.1101/2024.05.29.24308057

**Authors:** Philipp Wegner, Marcus Grobe-Einsler, Lara Reimer, Fabian Kahl, Berkan Koyak, Tim Elter, Alexander Lange, Okka Kimmich, Daniel Soub, Felix Hufschmidt, Sarah Bernsen, Mónica Ferreira, Thomas Klockgether, Jennifer Faber

## Abstract

Gait disturbances are the clinical hallmark of ataxia disorders, fundamentally impairing the mobility of ataxia patients. In clinical routine and research the severity of the gait disturbances is assessed within a well-established clinical scale and graded into categorial levels. Sensor-free motion registration and subsequent movement analysis allowed to overcome the obvious shortcoming of such coarse grading: Using time series models (tsfresh, ROCKET) we were not only able to successfully reproduce the categorial scaling (Human performance: 44.88% *F*_1_-score; our model: 80.28% *F*_1_-score). Particularly subtle, early gait disturbances and longitudinal progression below the perception threshold of the human examiner could be captured (Pearson’s correlation coefficient human performance -0.060, not significant; our model: -0.626, *p* < 0.01). Furthermore, SHAP analysis allowed to identify the most important features for each clinical level of gait deterioration. This could further improve the sensitivity to capture longitudinal changes tailored to the pre-existing level of gait disturbances (Pearson’s correlation coefficients up to -0.988, *p* < 0.01). In conclusion, the ML-based analysis could significantly improve the sensitivity in the assessment of gait disturbances in ataxia patients. Thus, it qualifies as a potential digital outcome parameter for early interventions, therapy monitoring, and home recordings.

## 1 Introduction

Neurodegenerative ataxias are a group of sporadic and hereditary movement disorders characterized by a progressive loss of balance and coordination accompanied by slurred speech leading to increasing disability and premature death. Gait disturbances are one of the clinical hallmarks resulting in substantial restriction of mobility with the need to use walking aids and finally the loss of ambulation in later disease stages. Initially, imbalance and resulting gait disturbances only become obvious during challenging tasks, such as a tandem walk or on uneven ground. As the disease progresses, also the normal gait becomes increasingly impaired. Since gait disturbances are one of the core features of ataxias, the clinical onset of the disease is most often defined as the patient’s reported onset of gait disturbances (Klockgether et al., 1998). The clinical scale for the assessment and rating of ataxia includes gait as the first item (Scale for the Assessment and Rating of Ataxia, SARA) (Schmitz-Hubsch et al., 2006). Here, a person’s gait is rated from 0 (normal) to 8 (unable to walk) based on two tasks: Task 1 is a 10m normal gait walk with a turn at the end followed by walking the same distance back. Task 2 is a walk in tandem (heels to toes) for at least 10 consecutive steps (Schmitz-Hubsch et al., 2006). The clinical rating is based on parameters such as missteps in tandem walk or staggering. For ataxia patients, alterations in e.g. step width while simultaneously decreased step length, increased variability in the placement and general trajectories of the feet have been described either from sensor-based gait assessments or with a multi-camera system. (Buckley et al., 2018; Ilg and Timmann, 2013; Kadirvelu et al., 2023; Seemann et al., 2024; Serrao and Conte, 2018). While this needs particular prerequisites, video-based markerless motion capturing has been used to study gait disturbances on a rodent model of ataxia (Lang et al., 2020). Lang et al. were able to characterize ataxia-specific movement in their model and further quantify subtle movement changes that could not be identified visually. In our work, we aimed to characterize gait disturbances in a large adult cohort of > 90 ataxia patients suffering from sporadic neurodegenerative or hereditary ataxias, as well as healthy controls autonomously by utilizing multiple machine-learning models within a straightforward sensor-free setting. For this purpose, we used participant’s normal gait that was videotaped during clinical examinations (*N* = 159). First, a deep learning-based markerless motion-capturing model was used to quantify a person’s gait by extracting time series of body markers and subsequently characterizing features thereof. Second, machine learning models were trained to reproduce the clinical classification, employing the human examiner’s SARA gait item scoring as ground truth. Third, we conducted a feature importance investigation to identify those features, most important for the final model prediction. Fourth, we analyzed the most important digital parameters in terms of their sensitivity to changes over time compared to the clinical scale. The aim of this study was to investigate to which extent machine learning (i) is suitable to reconstruct a clinical rating score usually determined by a trained neurologist, and (ii) can even improve sensitivity to detect subtle and longitudinal changes. Especially, in those hereditary ataxias, for which gene therapies are currently being tested in safety trials (clinicaltrials.gov NCT05822908), the detection of early subtle gait pathologies is of particular interest concerning their potential as digital outcome parameters in future prevention studies. The approach presented in this work offers great opportunities in assessing ataxia diseases more finegrained and personalized which ultimately allows improved disease modelling and prediction. In general, the digital assessment of gait disturbances based on markerless motion capturing is beneficial as it can be easily integrated into the clinical routine and could even be performed at home, allowing for closer monitoring of treatment responses and daily fluctuations.

## 2 Methods

### 2.1 Data

#### Participants & clinical assessment

119 participants of ongoing observational studies in neurodegenerative ataxias at the German Center for Neurodegenerative Diseases (DZNE) in Bonn, Germany, were included. Thereof were 91 ataxia patients suffering from various neurodegenerative ataxia disorders as follows: 58 spinocerebellar ataxia (SCA), 2 early-onset ataxias, 11 multiple system atrophy of cerebellar type (MSA-C) as well as 6 sporadic adult-onset ataxia (SAOA)), 3 FXTAS, 2 BRAT1, 2 SYNE1, 2 sporadic ataxias suspect for autoimmune ataxia, 2 Friedreich’s ataxia (FRDA), 1 CTX, 1RFC1 and 1 CANCA1A missense mutation. Moreover, 28 healthy controls (HC) were included in the study. All participants underwent a standardized clinical assessment including the Assessment and Rating of Ataxia (SARA) (Schmitz-Hubsch et al., 2006), which was videotaped, and the Inventory of Non-Ataxia Signs (INAS) (Jacobi et al., 2013). The first SARA item ‘gait’ consists of two tasks: (i) normal gait: normal gait walk for a 10m distance with a turn at the end on point followed by walking the same distance back (Task 1) and (ii) tandem walk: walk in tandem (heels to toes) for at least 10 consecutive steps (Task 2) (Schmitz-Hubsch et al., 2006). Graded scaling of the SARA gait item includes the assessment of both tasks and ranges from 0 (normal) to 8 (unable to walk even with assistance). The study was approved by the local ethics committee and all patients gave written consent including the assessments and the videotaping according to the Declaration of Helsinki.

### 2.2 Human rating

The on-site assessment of both SARA gait item tasks of normal gait and tandem walk instructed and rated by a certified rater was considered as ground truth for the SARA gait score. For the clinical rating by the human examiner, the evaluation of both tasks is needed. Particularly, the tandem walk needs to be considered for the discrimination between SARA gait scores 0 and 1. An unimpaired, normal gait with no difficulties in walking and turning as well as an unimpaired tandem walk is rated with 0, while the combination of an unimpaired normal gait but slight difficulties only visible when walking 10 consecutive steps in tandem is rated with 1. A subset of video recordings has been rated *a posteriori* by three clinical experts in consensus as part of the SARA training tool development (Grobe-Einsler et al., 2024) (N = 44). These posterior ratings were considered a human prediction and compared with the ground truth (onsite ratings). The resulting performance scores were taken as baseline performance for this work.

#### Video taping for markerless motion registration & selection of videos

In a total of 246 visits, SARA assessments were videotaped following a standardized protocol. For for the gait item, the camera is positioned directly in front of the proband (See Supplementary Figure S1). Notably, for the subsequent analysis of automated motion capturing, we only used the videos of Task 1, normal gait, of the SARA gait item. For quality control, all videos were inspected visually for suitability and excluded if unsuitable. Reasons for exclusion were: disturbances of the overall scene by other persons (e.g. clinic personnel), insufficient video quality, or environmental factors, such as mirroring surfaces, that disturb the motion capturing. We restricted the analysis to those patients who were able to perform the normal gait task without walking aids, which corresponds to a SARA gait score of less than 5. Thus, the final dataset included 159 recordings. In the end, each video is labeled with its respective on-site SARA gait score and additionally tagged as HC or ataxia patient.

### 2.3 Motion capturing

We used Alpha Pose, a full-body pose estimation model to extract movement markers from joints and other parts of the human body (Fang et al., 2023). Alpha Pose was favored over other frameworks such as OpenPose (Cao et al., 2019; Cao et al., 2017; Simon et al., 2017; Wei et al., 2016) due to its ease-of-use and superior performance on benchmark datasets commonly used in pose tracking (Fang et al., 2023). The Alpha Pose pipeline works in a two-step process which comprises at first a person localization step, using YOLOV3 (Redmon and Farhadi, 2018) and Efficent-Det (Tan et al., 2020), and subsequently a pose estimation step, while the latter utilizes ResNet (He et al., 2015) as its backbone model. Using the pose estimation model, 17 motion markers were extracted corresponding to 17 body parts, e.g. left ankle or right hip (see Supplementary Table S2 for details). The data provided by Alpha Pose yielded the horizontal x- and vertical y-positions for each marker in each frame of the video. Further on, from the Alpha Pose output, 4 multivariate time series were extracted, where each frame was considered a time step. (1) The first time series was based on the raw x positions of the hips, wrists, and ankles (each left and right respectively), which were assembled to a 6-dimensional (2+2+2) time series from here on referred to as *X-pos*. (2) Furthermore, distances between pairs of markers were extracted and formed the 6-dimensional time series here referred to as *Dist*. The elements of *Dist* are the distances between the ankles, between the wrists as well as the distances between the left hip and left wrist, between the right hip and right wrist, and the distance between the neck and left and right hip, respectively. (3) & (4) Finally, triples of markers were used to form triangles and extract angles from those at certain body positions. Those are from now on referred to as *Upper* and *Lower. Upper* consists of the angles at the shoulders, formed by the triangle shoulder, wrist, and hip, left and right respectively. *Lower* comprised the angles at the hips, formed by the triangle hip, left ankle, and right ankle, left and right hip respectively. Thus, two 2-dimensional time series resulted from that. In other words, each of the markers provided by Alpha Pose gives a certain value at a certain time step, respectively video frame. Observing these values, or values derived from those (e.g. angles and distances), over time yields a time series. This data generation, is further illustrated in Supplementary Figures S6, S7, S9, and S8.

### 2.4 Models and training

Two approaches, aimed to solve the task of SARA gait score reconstruction, were implemented, with each approach consisting of a classifier and a regressor. In the first approach, we combined tsfresh (Christ et al., 2018) and XGBoost (Chen and Guestrin, 2016). Here, time series features were obtained using tsfresh, which is a Python package that allows the extraction of 794 time series features from a single 1-dimensional time series. For the categories of multivariate time series as used in this work (*X-pos, Dist, Upper, Lower*), one obtains N*794 features, with N being the dimensionality of the times series included in the respective category (6, 6, 2, 2). The 794 time series features are predefined (Christ et al., 2018) and include parameters such as the variance of all values or the number of local maxima for each time series. To perform the final predictions a XGBoost model - classifier and regressor - was fitted on a subset of these extracted features to perform the final prediction. This subset of features resulted from a feature reduction procedure described further down.

The second approach utilizes ROCKET, a time series model that was able to report state-of-the-art performance scores in several time series classification tasks with significantly lower computational costs than established methods (Dempster et al., 2020). ROCKET extracts time series features by applying random convolutional kernels to the time series. Subsequently, a linear model is fitted on these features to perform the final prediction. As in (Dempster et al., 2020), ridge regression was used as the linear model and 10,000 was chosen as the number of convolutional kernels. The regularization strength used for the ridge regression was optimized in an inner cross-validation.

Both models, tsfresh and ROCKET, were fitted on the time series *X-pos, Dist, Upper*, and *Lower* as well as every combination of those. Each result reported in the following results section states the best score reached among all of these time series (-combinations). For instance, a model may achieve a worse performance using *X-pos* alone instead of using *Dist* and *Upper* combined. Note that when combining two multivariate time series of dimension *n* and *m* the resulting time series in a multivariate time series of dimension *n*+ *m*, e.g. *X-pos*+*Dist* is a 12 (6+6) dimensional time series.

As mentioned above, both models were trained and evaluated in a regression and classification scenario. In the regression setting, the models were trained to predict on the SARA gait score range [0,4] in all participants while the classification problem was subdivided into 15 binary classification problems, with one binary classification for every pair out of the SARA gait scores [HC,0,1,2,3,4], excluding self-pairs. Note that in the classification scenario, the ratings are considered as classes rather than values on an ordinal scale, and healthy controls were considered as their own class. Thus, within the classes of SARA gait scores 0-4 comprised ataxia data only. The regression problem is evaluated in terms of Root Mean Squared Error (RMSE) and *R*^2^-score. To test whether the models could distinguish between neighboring classes within the regression scenario, a Mann-Whitney U-test was conducted on the predicted values of neighboring classes and statistically significant differences between neighbors were reported. Additionally, the regression experiments are repeated excluding HC from the data. To account for class imbalances we report the classification scores in terms of a macro-averaged *F*_1_-score. That score is formed by calculating the *F*_1_-score for each class and averaging the result.

All models were trained and evaluated in a leave-one-out cross-validation with a slight modification as follows: For those participants (N = 30) with follow-up visits, all other videos of the participants whose video is currently in the test set were excluded. This was undertaken to avoid shortcut learning theoretically possible by the model recognizing a certain participant by personal gait characteristics. Additionally, the cross-validation of the tsfresh models included an inner cross-validation in which the number of features was reduced to the top k features ranked by SHAP (SHapley Additive exPlanations) values. A feature reduction mechanism was employed since the feature vectors assembled from tsfresh are high-dimensional and reducing the dimensionality in a sophisticated way can remove redundant data and avoid overfitting (Khalid et al., 2014; Mwangi et al., 2014). The feature selection mechanism used in this analysis has been shown to provide better results in feature reduction than other commonly used strategies (Marcilio and Eler, 2020). The parameter k was tuned in the inner loop with a 5-fold cross-validation on the current training set and accordingly can differ in each leave-out setting. Subsequently, the calculated SHAP values are used in terms of model explainability. Incorporating SHAP for model explainability has become a popular choice in machine learning (Fryer et al., 2021) and hence was also used in this work. The SHAP values were accumulated during the entire training process of the tsfresh models and finally, the extracted time series features were ranked by their respective SHAP values. The mean SHAP values are interpreted as feature importance and allow us to gain insight into the final model prediction. The regression experiments produce predictions on a floating scale which is not directly comparable to the integer-valued SARA gait scores resulting from the human baseline effort. However, a crucial step to investigate whether the proposed models are capable of tackling the task of rating gait disturbances according to SARA was necessary to compare their predictive performance to the human baseline performance. To map the float-valued predictions of the regression models to the actual SARA gait scale, the following procedure was undertaken. For one assessment, the regression model was used to locate the rating on the floating SARA gait scale. Subsequently, a specific classification model was incorporated to determine the final class. For instance, the regression model predicted a score of 2.42, subsequently a classification model trained on distinguishing SARA gait scores 2 and 3 was used to determine whether the final SARA gait score is a 2 or a 3. This was done for both tsfresh and ROCKET. Finally, the resulting integer-valued predictions were used to compare the models of this work with the human baseline.

### 2.5 Longitudinal analysis

Thirty patients were followed up longitudinally, which allowed to investigate the performance of the extracted gait features in capturing progressing gait disturbances that are expected over time compared to the clinical scale. For each feature extracted from the time series utilizing the tsfresh package, a correlation analysis was performed that investigated for which feature the greatest Pearson’s correlation coefficient was found. The correlation coefficient was formed between the value of that feature and the time variable. All SARA gait scores assessed in these 30 patients were normalized with respect to their initial baseline visit and the variable was numerically handled in days since the baseline visit. Finally, a linear regression analysis revealed whether the linear correlation between feature values and days since the baseline visit was significant. This investigation was performed on the entire longitudinal cohort (N=30) and on four sub-cohorts that were derived from grouping patients with the same SARA gait score assessed during their baseline visit. For instance, one sub-cohort was formed by grouping patients with a SARA gait score of 2 during their baseline visit. Finally, this study considered the predictions of the two models, tsfresh and ROCKET, derived from the regression experiments as a variable, here referred to as floating point SARA gait score, and investigated for correlation with the days since the baseline visit in the same manner as with the time series features.

### 2.6 Fairness analysis

Since this work aims to be incorporated into clinical practice in the future, it was necessary to investigate whether the models resulting from this work are fair. Here, we consider the models as fair if no sex or age group has to expect a significantly lesser performance from it. Hence, the mean absolute error for males and females as well as for the age groups 19-39, 40-59, and 60-82 were calculated and reported.

## 3 Results

Table 1 summarizes the demographic data of the patient and human control cohorts. 159 videos of participants performing the 10m walk formed the basis for the analysis. 87 videos had to be excluded. The on-site ratings, which serve as the ground truth, were distributed as depicted in Supplementary Table S3.

**Table 1:**
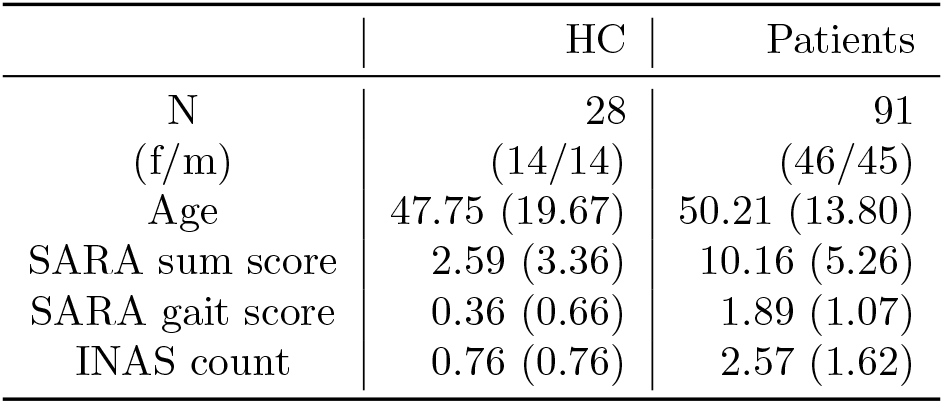
Demographic and characterizing data. Information is given as mean and standard deviation in brackets, resp. female (f)/male (m) distribution. INAS count was available for 139 and SARA sum score was available for 149 videos. Age, INAS, SARA, and SARA gait score is averaged over all videos within the respective group. The mean/standard deviation of SARA sum score, SARA gait score, and INAS count were calculated by including all visits of each participant.

The age distribution across the groups of ataxia patient subgroups according to their baseline SARA gait scores as well as the HC can be found in Supplementary Figure S5. The upcoming section presents the results of the two modeling approaches in both the regression and classification scenarios. Subsequently, this section presents the results of the explainability efforts and concludes with the results of the longitudinal analysis and fairness analysis. Note that in this section the models are referred to as tsfresh or ROCKET, however, tsfresh stands for the model using time series features generated from tsfresh and then fitting an XGBoost model on that. ROCKET refers to the model applying random kernels on the time series and subsequently fitting a ridge regression model on the result.

### 3.1 Human baseline

Out of the 159 videos, 44 were used to compare the on-site rating (the ground truth) with a posteriori consensus rating of the videos by three trained neurologists (Grobe-Einsler et al., 2024). The resulting human baseline performance scores are depicted in Table 3. The posterior consensus rating was able to reconstruct the SARA gait score with a macro-averaged *F*_1_-score of 44.88%, precision of 54.88%, and recall of 49.33%.

### 3.2 Regression Experiments

The evaluation of tsfresh and ROCKET prediction of the SARA gait score yielded RMSE and *R*^2^ scores presented in Figure 1. The experiments were conducted with every time series combination possible and the reported scores are the best noted, in terms of *RMSE* and *R*^2^-score. The tsfresh model, which utilizes explicit time series features, e.g. number of peaks, was able to score an *RMSE* of 0.824 while reporting an *R*^2^-score of 0.505. These scores were reached by utilizing the combination of times series *X-pos*+*Lower*, i.e. the body markers taken from the 6 raw x-positions combined with the two angles at the left and right hip of a person. Moreover, the model demonstrated a statistically significant (p < 0.05) distinction in the distribution of predictions between videos rated as 1 and those rated as 2. This suggests that the model effectively captured the nuances between these closely adjacent values on the SARA gait scale. Additionally, this was the case for the neighboring values 3 and 4. The model utilizing implicit time series features by applying random convolution kernels, ROCKET, was able to score a greater *RMSE* and *R*^2^-score than tsfresh was able to do. The ROCKET model was able to score an *RMSE* of 0.763 and an *R*^2^-score of 0.575. Furthermore, the statistical analysis revealed that the model predicted significant differences in SARA gait scores between videos rated as 0 and those rated as 1. This was also the case for two more neighboring scores, i.e. 1 and 2, and 2 and 3. The scores presented were scored by the ROCKET model adopting the time series combination *X-pos*+*Upper*+*Dist*, i.e. the 6 raw x-positions, the angles at the shoulders, and the 6 distances. All regression experiments were repeated excluding those participants without HC. Supplementary Figure S2 and S3. Table 2 depicts the results of the same experiments excluding HC together with the previously presented scores for comparison. The results for tsfresh show that including HC yielded a greater *R*^2^-score than excluding these. However, in terms of *RMSE* the opposite was observed. Including HC led to a higher *RMSE*, which is to be interpreted as a worse performance. Considering the ROCKET model, both scores, *RMSE* and *R*^2^-score, benefited from including HC. That means a greater *R*^2^-score while simultaneously reaching a lower *RMSE*.

**Table 2:** Regression results for different cases for handling HC. Results for tsfresh in the upper half of the table, ROCKET results in the bottom half of the table. Arrows indicate the favorable outcome, i.g. for *R*^2^ higher values are favorable, for RMSE lower values. X, D, U, and L indicate which time series combination lead to the best outcome. X=*X-pos*, D=*Dist*, U=*Upper*, L=*Lower*

| Case | Time Series | $R^2 \uparrow$ | RMSE ↓ |
| --- | --- | --- | --- |
| tsfresh |  |  |  |
| Including HC | X+L | **0.504** | 0.823 |
| Excluding HC | U+L | 0.482 | **0.769** |
| ROCKET |  |  |  |
| Including HC | X+D+U | **0.575** | **0.762** |
| Excluding HC | X+D+U | 0.470 | 0.778 |

**Figure 1:**
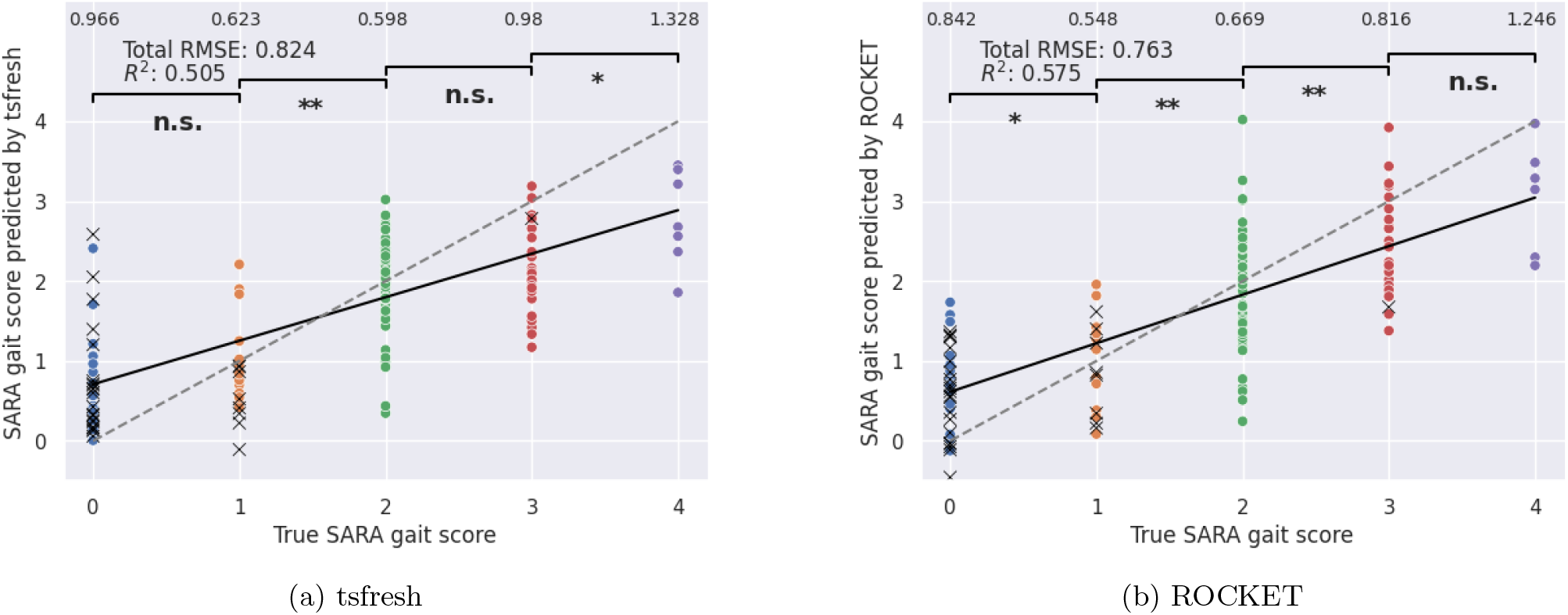
Regression experiments results. Results of tsfresh (left) and ROCKET (right) in terms of RMSE and *R*^2^-score. The numbers at the very top row present the RMSE constrained to that specific SARA gait score, e.g. the tsfresh model scored an RMSE of 0.623 on the SARA gait score 1. The thick black line is a linear fit on the model predictions while the dotted line is the diagonal representing a theoretical 1:1 relationship between true and predicted values. The brackets in the top section indicate whether the model predicted significant differences between neighboring SARA gait scores. Ataxia patients are represented by colored dots, HC are represented by black crosses. * *p* < 0.05, ** *p* < 0.01, not significant (n.s.)

### 3.3 Classification Experiments

#### 3.3.1 Binary Classification Experiments

The classification experiments comprised 15 experiments for each model architecture, tsfresh, and ROCKET. Figure 2 presents the results of these experiments in color-encoded matrices. Considering tsfresh (Figure 2(a)) a clear trend is observable where the greater the distance between two classes - in other words, the absolute difference between the two corresponding SARA gait scores - the more accurately the model could distinguish between them. The maximal *F*_1_-score with 95.38% was reached for the classification experiment separating SARA gait scores 0 and 4. The lowest *F*_1_-score with 60.92% came from the binary classification experiment that was trained on distinguishing SARA gait scores 0 and 1 of the patient group. Notably, tsfresh achieved an *F*_1_-score of 78.78% in the classification between HC and SARA gait score of 0 in the patient group. The tsfresh model utilizes explicit time series features and performed best when incorporating either the time series *Lower* or *Upper* or *Upper*+*Lower*, i.e. the two angles at the shoulders and two angles at the hips, in the majority of all binary classifications experiments (in 13 out of 15). Generally, the ROCKET model showed lower *F*_1_-scores for the binary classification experiments. As before, the more distant two classes were, the better the classification performance. The best performance was achieved by the ROCKET model with a perfect 100% *F*_1_-score in the classification of the SARA gait scores 1 versus 4. That score was achieved by incorporating the time series combination *X-pos*+*Dist*, namely the raw x positions of 6 markers combined with the 6 distances between markers. Here, the lowest reported *F*_1_-score of 55.85% was related to the classification of HC and SARA gait score of 1 in patients. Contrary to the tsfresh model, for the ROCKET model the most preferred time series are *X-pos* and *Dist*. All top-ranking *F*_1_-scores, in the binary classification experiments, were reported by utilizing either *X-pos* or *Dist* or both. On average, the tsfresh model could reach a macro-averaged *F*_1_-score of 83.65% (standard deviation (SD) 10.28) while ROCKET reached an *F*_1_-score of 76.70% (SD 13.89).

**Figure 2:**
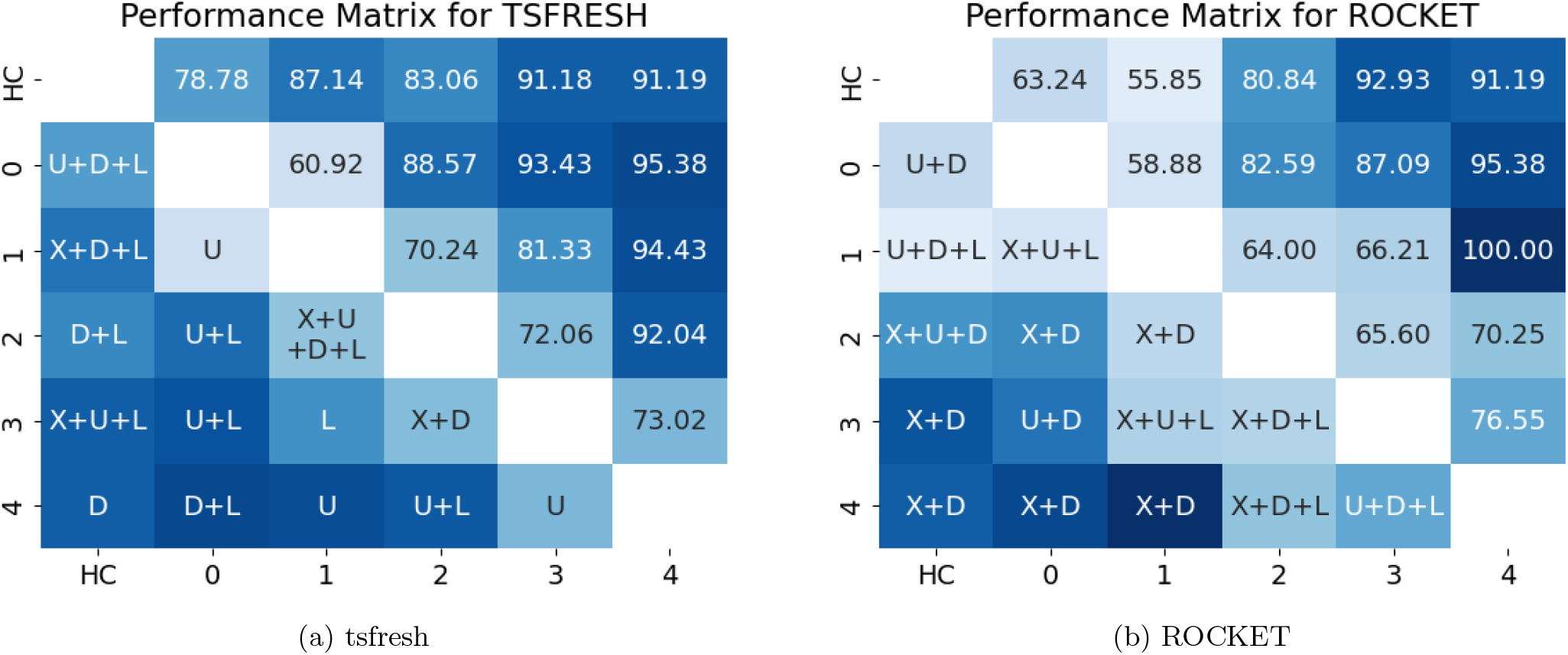
Classification experiments results. Results of tsfresh (a) and ROCKET (b) models. Each row-column combination in the upper right triangle depicts the best reported macro-averaged *F*_1_-score (in %) for the respective binary classification. The lower left triangle depicts for which time series or combination of time series this performance was reported. For instance, the best tsfresh model trained to classify between the SARA gait scores 0 and 1 within the ataxia group was able to score a macro-averaged *F*_1_-score of 60.92% when using the times series assembled from the angles at the shoulders only (*Upper*). X=*X-pos* (time series of raw x-positions of each marker separately), D=*Dist* (time series of distances between two markers), U=*Upper* (times series of angles of the upper body part, e.g. shoulders), and L=*Lower* (times series of angles of the lower body part, e.g. hip).

#### 3.3.2 Evaluating on the entire SARA gait range

Finally, this work evaluated the ability of tsfresh and ROCKET to classify between all 5 SARA gait classes considered, namely [0, 1, 2, 3, 4] (Table 3). HC was not considered as a separate group in this experiment. The presented scores are macro-average scores over all classes.

**Table 3:** Classification results on entire scale. Results of the classification experiment on the SARA gait score classes [0,1,2,3,4], presented in %. {ROCKET, tsfresh}_*H*44_ indicates that the models were evaluated on the subset of the 44 cases, which were used to create the human baseline. Both models outperformed the human baseline. All scores are presented in %. Prec.=Precision, Rec.=Recall, *F*_1_=*F*_1_-score. Arrows indicate the favorable outcome, for all considered metrics, higher values are favorable. n.a.=not applicable

| Model | Features | Prec. ↑ | Rec. ↑ | $F_1$ ↑ |
| --- | --- | --- | --- | --- |
| *tsfresh* | U+L | 57.92 | 50.21 | 52.39 |
| *tsfresh*<sub>H44</sub> | U | 63.77 | 54.31 | 52.74 |
| *ROCKET* | U+L | 85.16 | 78.59 | **81.23** |
| *ROCKET*<sub>H44</sub> | D+U+L | 88.80 | 76.98 | **80.28** |
| *HumanBaseline* | n.a. | 54.88 | 49.33 | 44.88 |

Both models outperformed the human baseline. Evaluating tsfresh on all samples achieved an *F*_1_-score of 52.39% and 52.74% when the data was restricted to the 44 assessments retrospectively rated by three neurologists (human baseline). ROCKET, when evaluated on all samples, achieved an *F*_1_-score of 81.23% and 80.28% when the data was restricted to the 44 samples of the human baseline. Thus, here the model based on random convolution kernels, ROCKET, yields superior performance compared to tsfresh and the human baseline.

### 3.4 Explainability

Since the implementation of tsfresh features involved a feature reduction mechanism that uses SHAP values, we were able to utilize these values to gain insight into feature importance. Accumulating these SHAP values for each time series feature over the entire training process allowed us to evaluate which markers were most important for the model to generate its final prediction. Figure 3 presents the results of this investigation in a radar plot. The most important features originated from the time series comprising the angles of the upper and lower body. The angles at the left shoulder achieved the highest SHAP value on average, followed by those of the right shoulder. Features derived from the angles at the hips were on rank 4 (right hip) and 6 (left hip). In between, on rank 5 was the first distance feature: distance between both ankles. Lesser importance was given to those features taken from the raw x positions of certain body markers (*X-pos*) and from the pairwise distances (*Dist*), except for the distance between the ankles. The same analysis was performed for neighboring classes in the classification experiments. Neighboring classes here refer to the binary classifications HC vs. 0, 0 vs. 1, 1 vs. 2, 2 vs. 3, and 3 vs. 4. This is presented in the Supplementary Table S4. The pairwise analysis revealed that for the model to distinguish between the HC and SARA gait scores 0 in the ataxia group as well as to distinguish between 0 and 1, the most important body marker is the angle at the right shoulder. Concerning 1 vs. 2, the distance between the neck and the left hip received the greatest SHAP value. Moving on to 2 vs. 3, the distance between the ankles and the distance between the neck and right hip share a similar high SHAP value. Finally, to separate the SARA gait scores 3 and 4, the model benefited the most again from the time series features extracted from the angle at the right shoulder. Summarizing the explainability analysis of the binary experiments yielded that the markers on the right side of the body tend to receive higher feature importance than the markers on the left side, and particularly the angles exhibit the highest feature importance.

**Figure 3:**
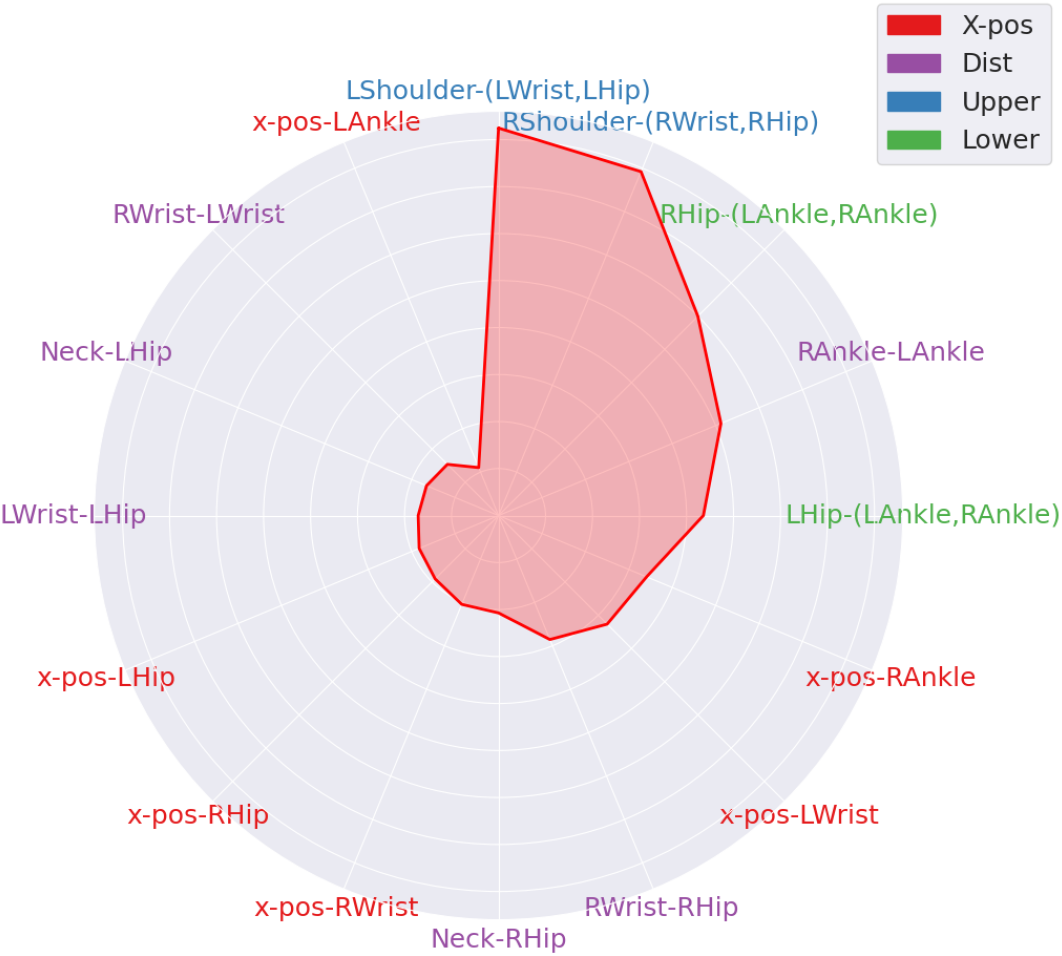
Explainability analysis. Mean SHAP values by marker presented in a radar plot. SHAP values are calculated during the feature reduction step implemented in the regression experiment of the pipeline utilizing the tsfresh model.

### 3.5 Longitudinal analysis

Thirty participants returned for at least one follow-up visit. Table 4 characterizes this longitudinal sub-cohort. To investigate features suitable to model gait disturbances in ataxia over time, we plotted the relative change to the value assessed at the baseline visit of each considered parameter against the time since baseline in days 4. This section refers to particular features, e.g. numbers of local minima, of a respective time series, e.g. x position of the left hip (as part of the *X-pos* time series) or the angles of the right shoulder (as part of the *Upper* time series). Thus, a longitudinal deterioration of gait disturbances might result in either a decrease or an increase, e.g. negative or positive deltas compared to baseline. Detailed information on the selected features is given in Supplementary Table S1.

**Table 4:** Characterizing data of the longitudinal sub-cohort. The baseline rating refers to the SARA gait score rating assessed by a neurologist during the first SARA assessment of the respective participant. 24 patients had 1 follow-up visit, 6 patients had 2 follow-up visits.

|  |  |
| --- | --- |
| Time of observation in days<br>mean, [min, max] | 463.43 [130, 810] |
| Number of visits: | 2 visits: 24<br>3 visits: 6 |
| Number of patients<br>according to their baseline<br>level of gait disturbances: | SARA gait score of 0: 5<br>SARA gait score of 1: 4<br>SARA gait score of 2: 15<br>SARA gait score of 3: 5<br>SARA gait score of 4: 0 |

#### Clinical scale

The clinical scale, SARA gait score, itself failed to show any significant longitudinal changes in the studied cohort. Increases and decreases relative to the SARA gait score at baseline were roughly balanced, whereby the expected deterioration in gait was barely captured with a Pearson’s correlation coefficient of -0.06.

#### Overall time series feature

For the overall analysis of the whole ataxia cohort the x-position of the left hip was identified as the time series containing the one feature best modeling the longitudinal change. The correlation of feature change to baseline versus days since baseline was significant (Pearson’s correlation coefficient -0.625, p<0.01) (Figure 4 B).

**Figure 4:**
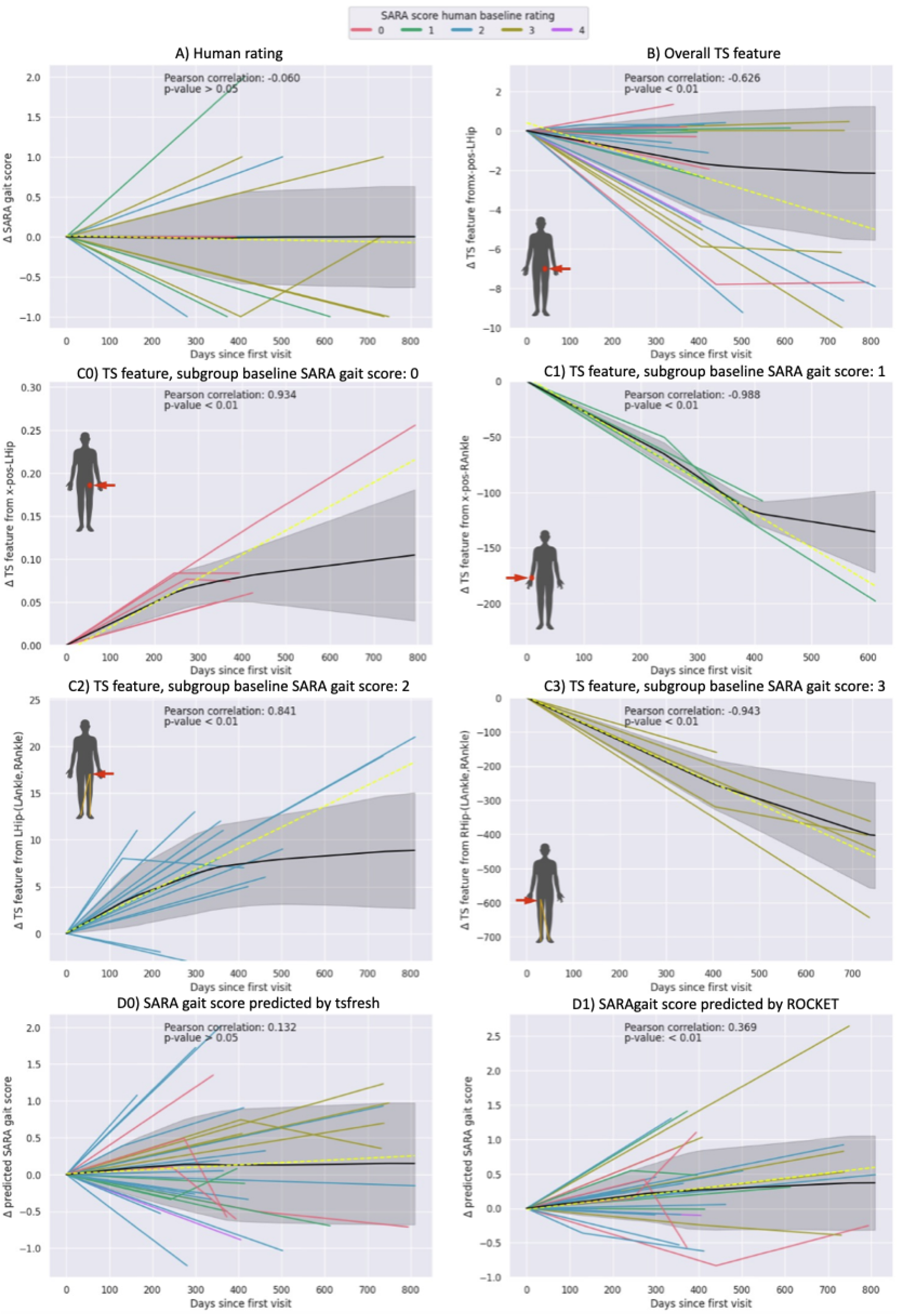
Longitudinal analysis. The relative change to baseline of the on-site SARA gait score (human baseline, ground truth) (A), the overall (B) as well as stage-dependent particular features of certain time series (C0-C3) and the floating point SARA gait scores predicted in the regression task of each model, tsfresh (D0) and ROCKET (D1), are plotted against the time since baseline in days. The stage-dependent features were studied in subgroups of respective ataxic gait severity levels at baseline: SARA gait score of 0 (C0), 1 (C1), 2 (C2), and 3 (C3) at baseline. The black line represents the mean trajectory, created by a linear interpolation, with the gray shaded area being the respective standard deviation. The yellow dotted line illustrates the linear regression fit of the given data. Pearson’s correlation coefficient and a p-value from the linear regression analysis are given. For the time series features (B, C0-C3) the anatomical miniature reference depicts the respective time series from which the feature was extracted. TS = time series, R = right, L = left

#### Stage-dependent time series features

Further stratification of the longitudinal cohort by their SARA gait score rating at baseline allowed to identify features tailored to the stage-dependent severity of ataxic gait. All stage-dependent time series features were significant and reached higher correlation coefficients than the overall time series feature (3.5) ranging from absolute Pearson’s coefficients between 0.841 to 0.988 compared to 0.625. For patients that started with a baseline SARA gait score rating of 0, again one feature of the time series constructed from the x position of the left hip was identified as the best feature for modeling longitudinal changes (Pearson’s coefficient of 0.934, p<0.01)(Figure 4 C0). For patients with a baseline SARA gait score rating of 1 a certain time series feature of the x position of the right ankle best modeled the longitudinal change with the highest absolute observed Pearson’s coefficient of -0.988 (p<0.01)(Figure 4 C1). For patients initially rated with a SARA gait score of 2 a feature of the time series from the angle at the left hip (*Lower*) best captured the longitudinal change with a Pearson’s coefficient of 0.841 (p<0.01)4 C2). Finally, we considered those patients who were rated with a SARA gait score of 3 at baseline. For those, a feature from the time series of the angle at the right hip (*Lower*) best captured the longitudinal change yielding a Pearson’s coefficient of -0.943 (p< 0.01)4 C3). For patients with a SARA gait score of 4 only baseline assessments were available.

#### Predicted SARA gait score

The two final analyses performed on the longitudinal cohort aimed to answer whether the floating point SARA gait score, which was predicted by the two models, tsfresh and ROCKET (3.2) during the regression experiments, can capture a significant longitudinal change. Concerning the tsfresh model, this was not the case. A positive Pearson’s coefficient was reported with a value of 0.132 but this relationship was not significant (p>0.05). However, the model utilizing random convolutional kernels, ROCKET, predicted significantly increasing SARA gait scores over time with a Pearson’s correlation coefficient of 0.369 (p<0.01) (Figure 4 D0, D1).

### 3.6 Fairness

Evaluating the mean absolute error (MAE) produced by each model, tsfresh and ROCKET, on different age and sex groups gives insight into the fairness of the model. The model using ROCKET performs best applied to the age group 60 to 82 (MAE: 0.555) and worst for participants aged 40 to 59 (MAE: 0.707). The age group 19 to 39 ranked between the two other groups (MAE: 0.660). The tsfresh model produces residuals that yielded a similar MAE for all age groups (MAE: 0.665 for age 19-39; MAE: 0.657 for age 40-59; MAE: 0.637 for age 60-82). Concerning the split male/female, the ROCKET model produced an MAE of 0.579 for males and 0.645 for females. The tsfresh model on the other side produced an MAE of 0.702 for males and 0.556 for females. The results are presented in Table 5. In summary, no model showed relevant performance increases or decreases in any sex and/or age group.

**Table 5:** Fairness evaluation. Mean absolute error (MAE) of each model for the age and sex groups. MAE is given as mean and standard deviation (SD) in brackets.

|  | ROCKET<br>MAE | tsfresh<br>MAE |
| --- | --- | --- |
| Age-range / N |  |  |
| 19-39/35 | 0.660 (0.43) | 0.665 (0.55) |
| 40-59/59 | 0.707 (0.49) | 0.657 (0.53) |
| 60-82/34 | 0.555 (0.51) | 0.637 (0.41) |
| Sex / N |  |  |
| male/59 | 0.579 (0.45) | 0.702 (0.53) |
| female/60 | 0.645 (0.47) | 0.556 (0.44) |

## 4 Discussion

We used videotaped 10m walks of normal gait in a large cohort of ataxia patients and healthy controls and incorporated multiple machine learning methods to reproduce the human clinical rating of gait disturbances as well as for the assessment of longitudinal changes. A markerless motion-capturing model, AlphaPose, creates the time series by extracting the xy-position for every marker in every frame of the video. Two modeling approaches were used. In the first approach, referred to as tsfresh, a time series feature vector was generated using tsfresh and we subsequently fitted an XGBoost model on it. The second approach, ROCKET, applied random convolutional kernels to the time series and fitted a ridge regression on the generated features. Both models outperformed the human rater in capturing early, subtle gait disturbances as well as longitudinal changes.

The general performance of both models is convincing. The predicted (floating point) SARA gait values of both models showed a strong correlation with the human SARA gait rating (*R*^2^ > 0.5). As expected, the binary classification improved on average for more distant classes, e.g. greater difference between the two considered SARA gait scores. Moreover, there are two notable details, with regard to the superiority of the automated gait analysis to detect subtle changes occurring in the early disease course. First, within the classification experiments, the binary classification between HC and ataxia patients rated with a SARA gait score of 0 (normal gait) by the human examiner achieved higher macro averaged *F*_1_-scores than between human SARA gait score rating of 0 and 1 in ataxia patients. Moreover, the *R*^2^-score was slightly stronger in the entire cohort comprised of ataxia patients and HC than for the ataxia patients only, for both models. Thus, including HC might overall reduce variances within the group of participants rated with a SARA gait score of 0. We hypothesize, that both observations are related to the fact that ataxia patients, even if rated as 0 (normal gait) by the human examiner, might nonetheless already exhibit very subtle alterations or higher variability within their gait parameters. From a conceptual point of view a gradual deterioration is reasonable and particularly for hereditary ataxias, alterations of gait parameters - assessed with body-worn sensors or a 6-camera system - have been described for early, so-called pre-ataxic disease stages with an overall SARA sum scores < 3 (Ilg et al., 2022; Shah et al., 2021). Second, both models “outperformed” the human rater in the discrimination accuracy of SARA gait score 0 (“normal gait”) (Schmitz-Hubsch et al., 2006) and SARA gait score 1 (“slight difficulties only visible when walking 10 consecutive steps in tandem”) (Schmitz-Hubsch et al., 2006). In our video analysis the second task of the gait assessment, the tandem walk, was not considered. Thus, based on the normal gait task alone, the human rater should not be able to discriminate SARA gait scores of 0 versus 1. In contrast, the macro averaged *F*_1_-score of this particular binary classification was around 60% for both models, which again underlines the potential for the detection of subtle, early gait disturbances.

Incorporating machine learning models into clinical practice benefits from models being able to reason about their decisions. This not only allows clinicians to gain insight into the importance of certain features leading to the models’ final decision but also increases the trust in these models (Shin, 2021). Hence, this work included an explainability approach where the captured body markers are ranked by SHAP values. This analysis could only be carried out for the tsfresh approach, as this works with explicit time series features, in contrast to ROCKET with its random kernel approach. This “explainable AI” analysis revealed, that for the overall prediction of SARA gait scores, the top 5 features with the greatest influence on the model’s final prediction were features extracted from the time series of the angles at the shoulders and hips as well as the distance between both ankles. This seems reasonable since the ataxic gait is characterized by a widened base, staggering, and truncal instability with balancing movements of the arms. We also included the explainable AI approach for the longitudinal analysis. For the overall evaluation of longitudinal changes, a feature from the x-position time series of the left hip did by far outperform the human rating, which was not able to capture the expected longitudinal change, as well as the predicted floating point SARA gait scores of both models. Obviously, but nevertheless important to note is, that the selection of stage-dependent features tailored to the respective severity level of a participant further improved the detectability of progressive longitudinal changes. This is in line with previous studies in SCA2 with body-worn sensors (Seemann et al., 2024). In addition to the superiority in detecting subtle and longitudinal changes, the automated analysis was more accurate in the reproduction of onsite ratings compared to a human baseline. We have defined the on-site SARA rating of an experienced human examiner as ground truth. The clinical scale, SARA, has been shown to have a high inter-rater reliability. However, in comparison to both models, the reproduction of the on-site rating based on a consensus rating of the videos by three neurologists (referred to as human baseline) did show a lower *F*_1_-score than both models (44.88% vs. 52.74% for tsfresh and 80.28% for ROCKET). This might be due to the limited perspective with only the front video caption, while the human examiner on site has the advantage of the best positioning for them to rate the gait of participants. Yet, the possibility of bias on the part of the on-site investigator due to the general impression gained during the entire clinical visit cannot be ruled out. Nevertheless, this underlines the potential of digital methods to reproduce clinical scores.

With regard to the overall prediction of the SARA gait score, ROCKET showed a better performance with lower values for RMSE and a higher *R*^2^. One big advantage of the ROCKET model is its low computational costs. This would even make it possible to run ROCKET locally on a smartphone device, providing an elegant way to analyze gait parameters without the need to transfer video data, which can be favorable in terms of data protection. However, with regard to the discrimination of neighboring classes as well as for the identification of the most important features, tsfresh is more convincing. In particular, longitudinal changes were accurately recorded, when considering the pre-existing ataxia severity. Thus, allowing a very tailored monitoring of progression as desired within the framework of clinical trials. Moreover, since tsfresh still works with concrete time series features (instead of random kernels as ROCKET does), the explainability AI approach allowed to gain insights into the most important features overall as well as for particular patient subgroups. The test setup was very simple with a single camera positioned in front of the participant’s walking distance and is therefore suitable for fast and easy recording during clinical routine as well as for home recordings. Studies with recordings at home using a walking distance shorter than 10 m have already been demonstrated to have a good correlation with the established distance investigated here Grobe-Einsler et al., 2021. Interestingly, this study was also able to document the daily form-dependent fluctuations often reported by ataxia patients. The digital methods presented in our work allow large amounts of data to be analysed in a very short time, making it possible to evaluate high numbers of recordings. Therefore, they have enormous potential for overcoming the dependence on individual assessments in the clinical setting - which may be influenced by the current form of the day - as well as for a regular therapy monitoring at home.

## 5 Conclusion and Outlook

Applying machine learning to clinically assessed video data of normal gait using markerless motion capturing and fitting time series models on the results, as done in this study, has been demonstrated to be suitable for an automated rating of ataxic gait disturbances. Particularly, subtle early and longitudinal changes, not observable by a human examiner could be detected. Thus, it provides a feasible and easy-to-use tool for clinical routine as well as assessments at home. However, further investigation, including more data, particularly more longitudinal data in homogeneous disease groups, is needed to build more accurate models. In addition, comparative studies with wearable sensors are desired, as is the consideration of adding further recording angles.

## 6 Author contributions

MGE, JF, TK, and BK conducted the clinical part of this study. This comprised patient care and performing the SARA assessments. TE, AL, and PW gathered clinical information about the study participants. LR, FK, and MR consulted this work and contributed to conceptualizing the project. TK and JF supervised this work and contributed to the final paper writing. PW implemented the models, performed the analysis, and wrote the paper.

## Data Availability

All data produced in the present study are available upon reasonable request to the authors

## 7 Acknowledgements

This study was funded by the iBehave Network, sponsored by the Ministry of Culture and Science of the State of North Rhine-Westphalia. JF received funding from the Advanced Clinician Scientist Programme (ACCENT, funding code 01EO2107). The ACCENT Program is funded by the German Federal Ministry of Education and Research (BMBF).

## 8 Abbreviations and introduced terms

RMSE: Root mean squared error
MAE: Mean absolute error
*X-pos*: Multivariate time series constructed from the x positions of single body joints, namely left and right wrist, ankle, and hip (Supplementary Figure S6)
*Dist*: Multivariate time series constructed from the pairwise distance between the ankles, left hip and left wrist, right hip and right wrist, as well as neck and hips (Supplementary Figure S7)
*Upper*: Multivariate time series constructed from the angles of the triangles formed by the shoulder, wrist, and hip, each left and right respectively (Supplementary Figure S8).
*Lower*: Multivariate time series constructed from the angles of the triangles formed by (left and right) hip and both ankles (Supplementary Figure S9)

## 9 Supplementary Material

### S9.1 Implementation details

#### tsfresh model

The time series features were generated with the tsfresh framework implemented in Python https://tsfresh.readthedocs.io/en/latest/. The XGBoost models were taken from the Python implementations of XGBoost https://xgboost.readthedocs.io/en/stable/. Hyperparameters were tuned using Optuna https://optuna.org/. SHAP values were calculated using the SHAP python implementation https://SHAP.readthedocs.io/en/latest/index.html.

#### ROCKET model

We used the ROCKET implementation provided as a part of sktime https://github.com/sktime/sktime.

### S9.2 Supplemantary Tables

**Table S1:**
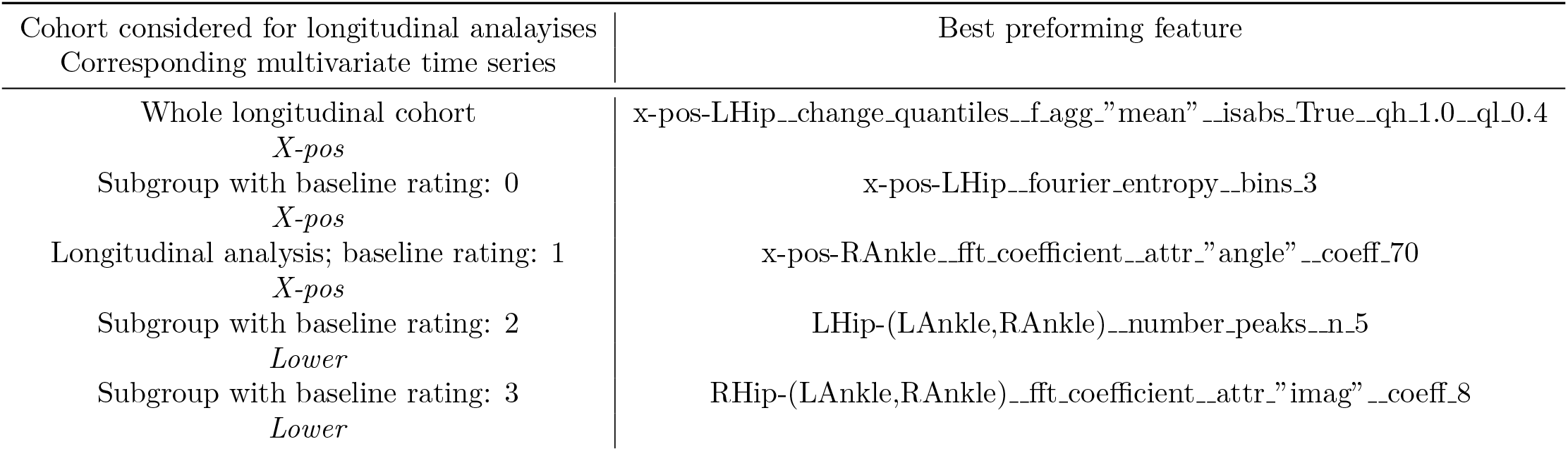
Time series features in longitudinal analysis. Time series features referred to in the longitudinal analysis 3.5, which did best model the longitudinal change. Note that implementations of the features can be found here: https://tsfresh.readthedocs.io/en/latest/text/list_of_features.html

| Cohort considered for longitudinal analyses<br>Corresponding multivariate time series | Best performing feature |
| --- | --- |
| Whole longitudinal cohort<br>*X-pos* | x-pos-LHip__change_quantiles__f_agg-"mean"__isabs-True__qh_1.0__ql_0.4 |
| Subgroup with baseline rating: 0<br>*X-pos* | x-pos-LHip__fourier_entropy__bins_3 |
| Longitudinal analysis; baseline rating: 1<br>*X-pos* | x-pos-RAnkle__fft_coefficient__attr-"angle"__coeff_70 |
| Subgroup with baseline rating: 2<br>*Lower* | LHip-(LAnkle,RAnkle)__number_peaks__n_5 |
| Subgroup with baseline rating: 3<br>*Lower* | RHip-(LAnkle,RAnkle)__fft_coefficient__attr-"imag"__coeff_8 |

**Table S2:** Body markers available. 17 body parts extracted by Alpha Pose model.

| Abbreviation | Internal Name | Body Part |
| --- | --- | --- |
| 0 | Nose | Nose |
| 1 | LEye | Left Eye |
| 2 | REye | Right Eye |
| 3 | LEar | Left Ear |
| 4 | REar | Right Ear |
| 5 | LShoulder | Left Shoulder |
| 6 | RShoulder | Right Shoulder |
| 7 | LElbow | Left Elbow |
| 8 | RElbow | Right Elbow |
| 9 | LWrist | Left Wrist |
| 10 | RWrist | Right Wrist |
| 11 | LHip | Left Hip |
| 12 | RHip | Right Hip |
| 13 | LKnee | Left Knee |
| 14 | RKnee | Right Knee |
| 15 | LAnkle | Left Ankle |
| 16 | RAnkle | Right Ankle |

**Table S3:**
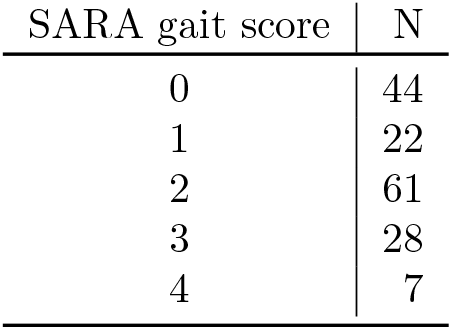
SARA gait score distribution. Distribution of SARA gait scores throughout the cohort used in this study.

| SARA gait score | N |
| --- | --- |
| 0 | 44 |
| 1 | 22 |
| 2 | 61 |
| 3 | 28 |
| 4 | 7 |

### S9.3 Supplementary Figures

**Figure S1:**
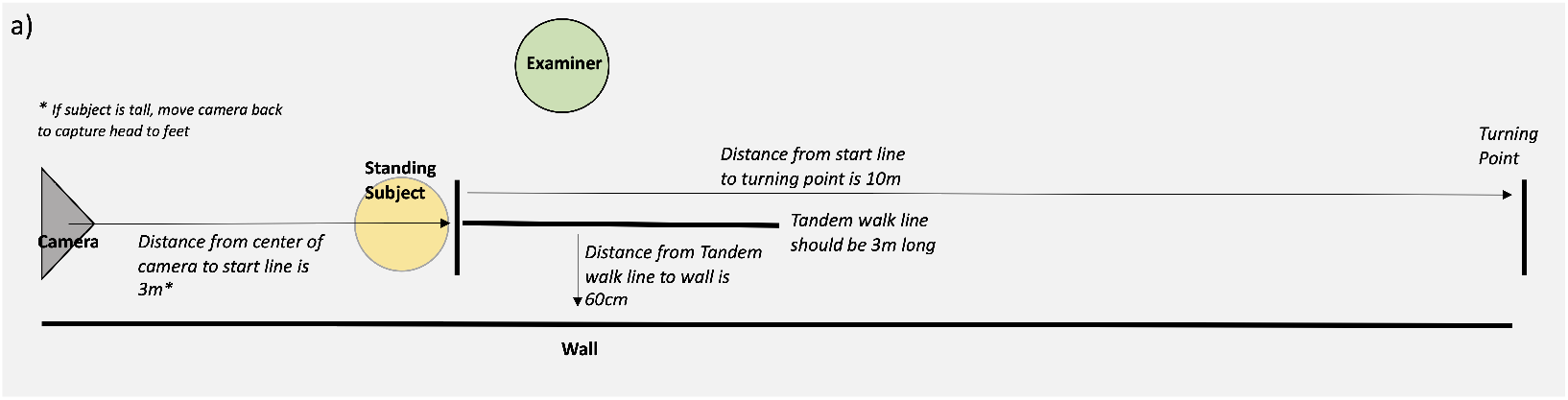
Video protocol. Protocol for the SARA video assessments. Figure from Grobe-Einsler et al., 2024.

**Figure S2:**
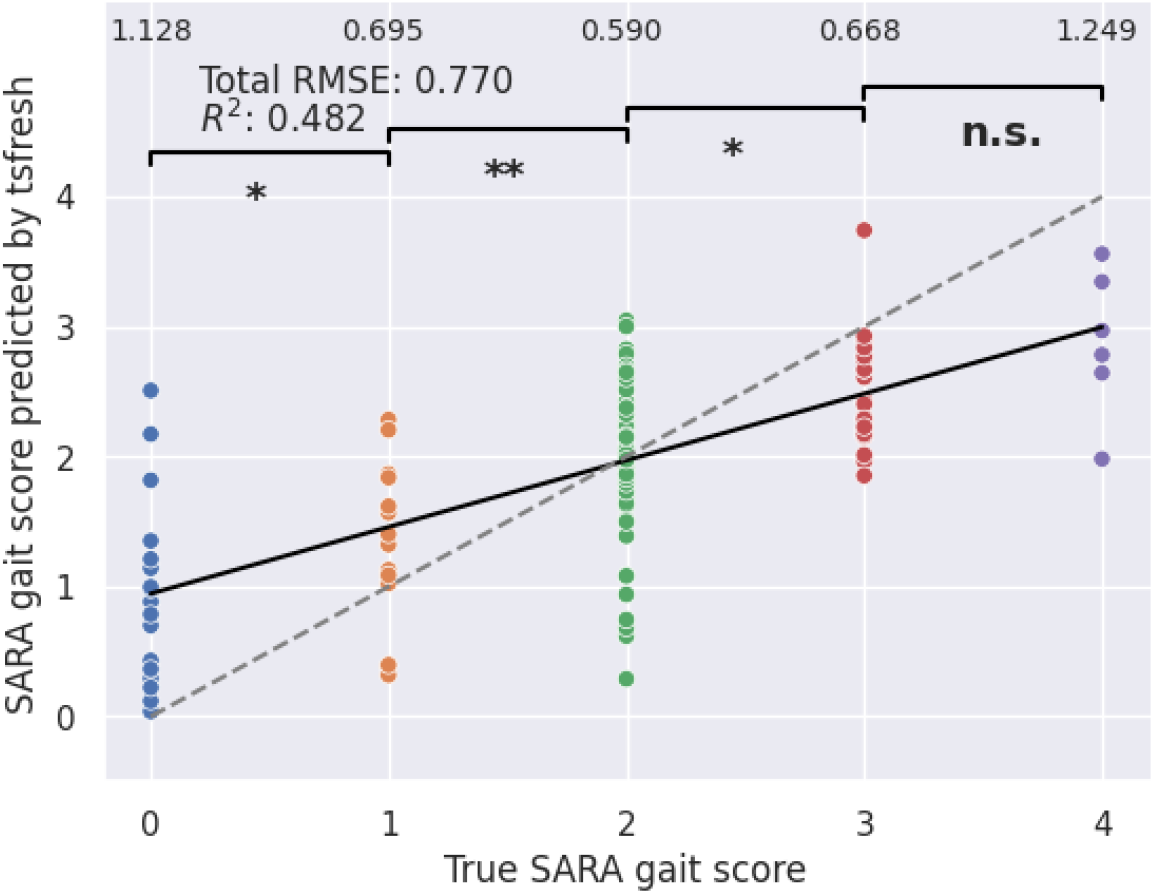
Regression results. Results of tsfresh in terms of RMSE and R2-score in the ataxia patient group only. The numbers above present the RMSE constrained to that specific SARA gait score, e.g. the model scored an RMSE of 0.590 on the SARA gait score 2. The thick black line is a linear fit on the model predictions while the dotted line is the diagonal representing a theoretical 1:1 relationship between true and predicted values. The brackets in the top section indicate whether the model predicted significant differences between neighboring SARA gait score scores. * *p* < 0.05, ** *p* < 0.01, n.s. = not significant.

**Figure S3:**
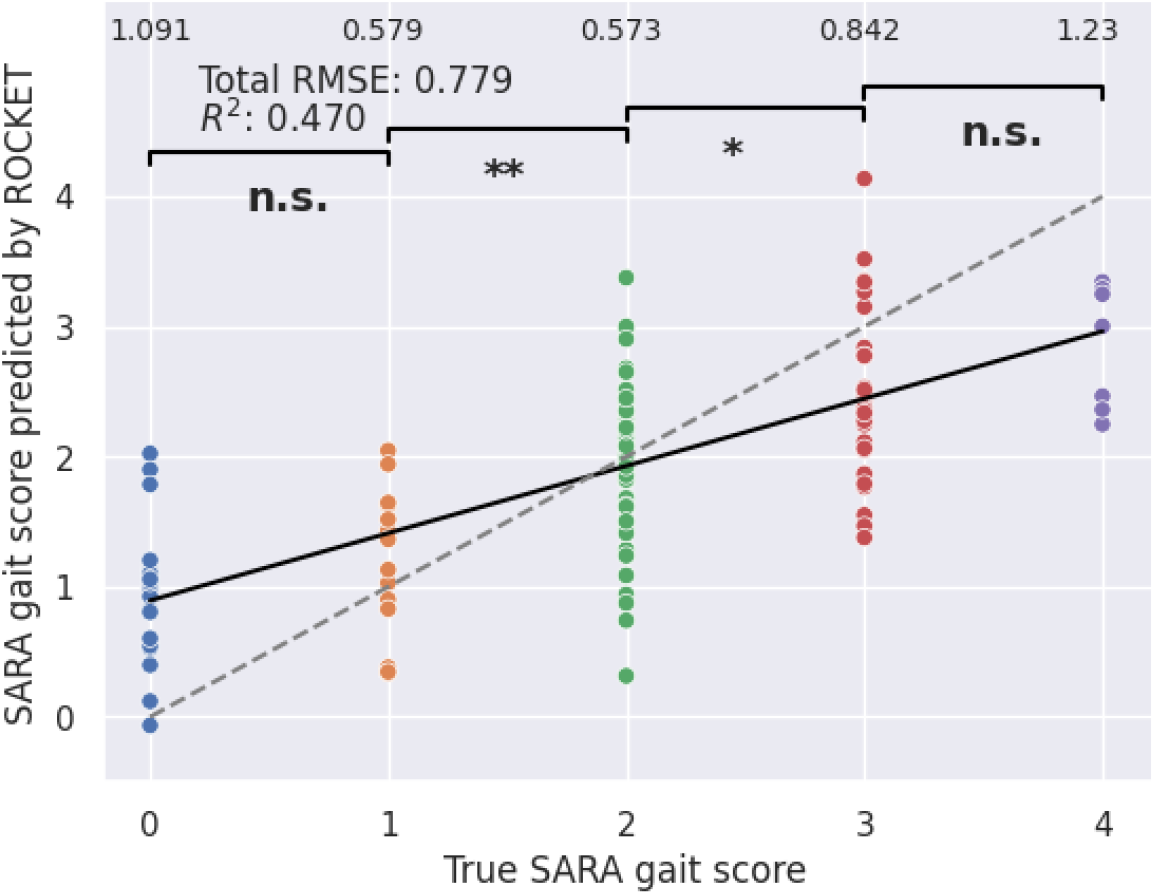
Regression results. Results of ROCKET in terms of RMSE and R2-score in the ataxia patient group only. The numbers above present the RMSE constrained to that specific SARA gait score, e.g. the model scored an RMSE of 0.573 on the SARA gait score 2. The thick black line is a linear fit on the model predictions while the dotted line is the diagonal representing a theoretical 1:1 relationship between true and predicted values. The brackets in the top section indicate whether the model predicted significant differences between neighboring SARA gait score scores. * *p* < 0.05, ** *p* < 0.01, n.s. = not significant.

**Figure S4:**
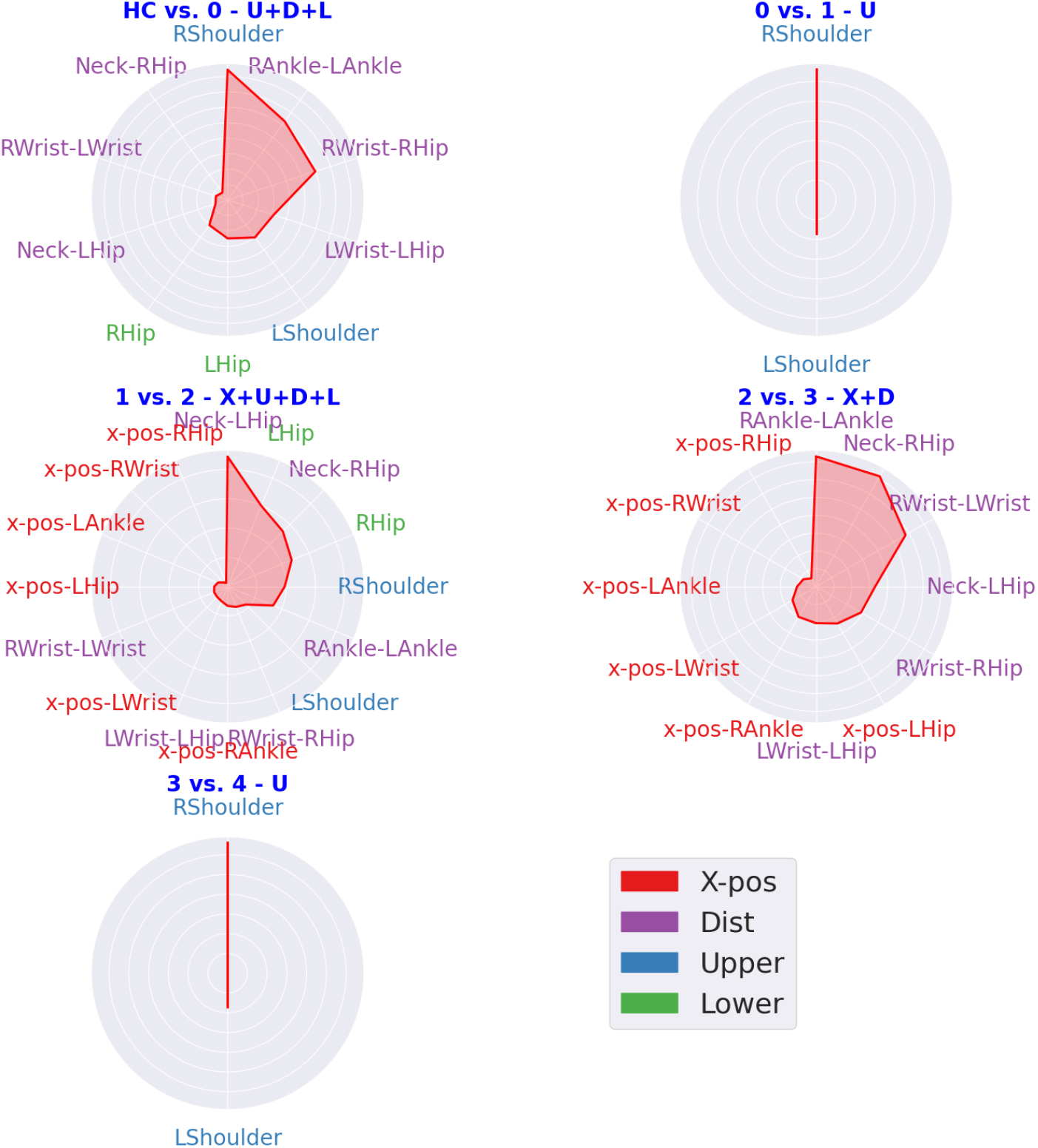
Explainability in classification experiments. Feature importance in direct neighbor comparisons in terms of SHAP values. Binary classifications of the neighboring classes: HC versus ataxia patients rated with a SARA gait score of 0; ataxia patients rated with a SARA gait score of 0 versus 1, etc. See Section 3.4

**Figure S5:**
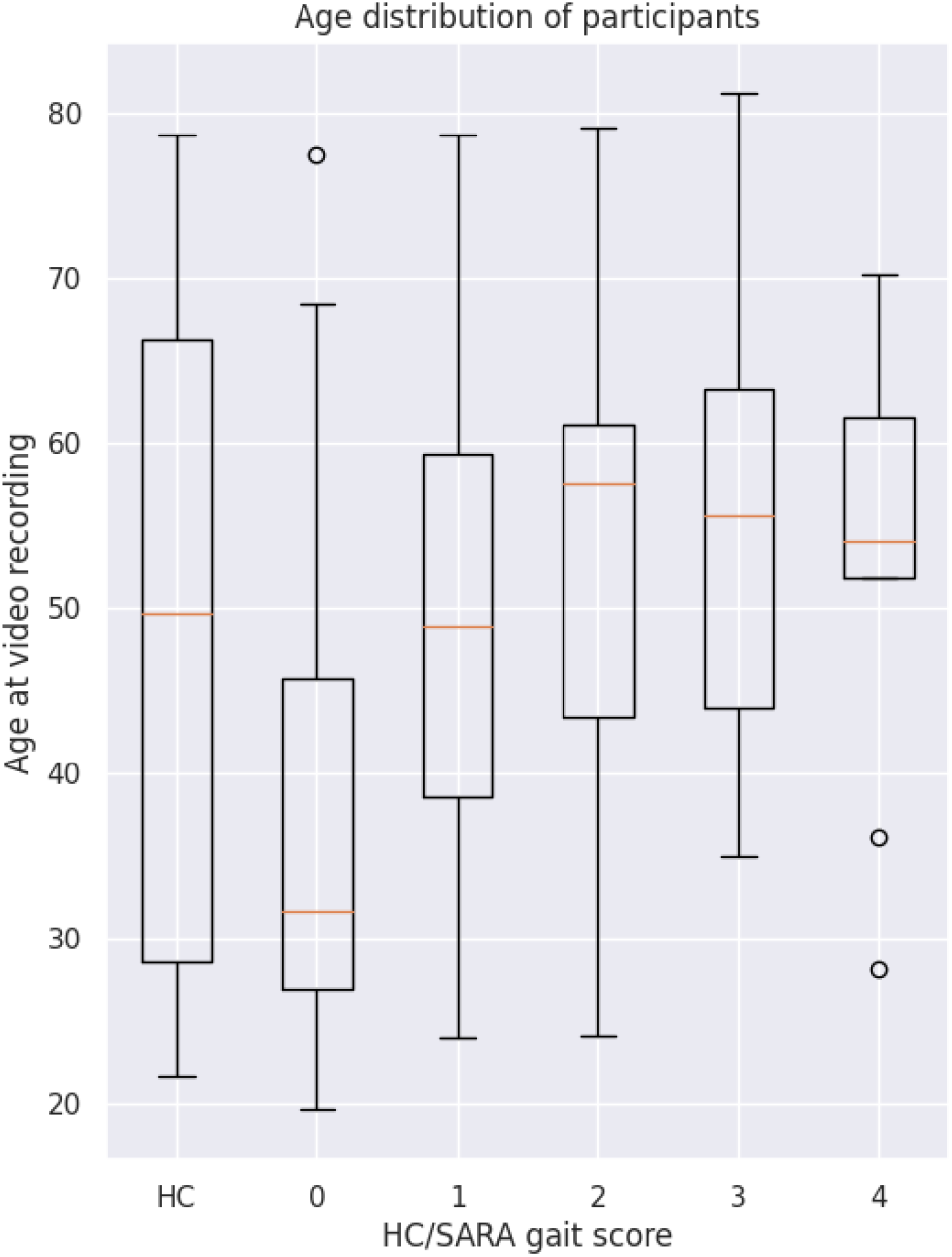
Age distributions. Age distributions of participants stratified by the subgroups of HC as well as for ataxia patients by SARA gait score.

**Figure S6:**
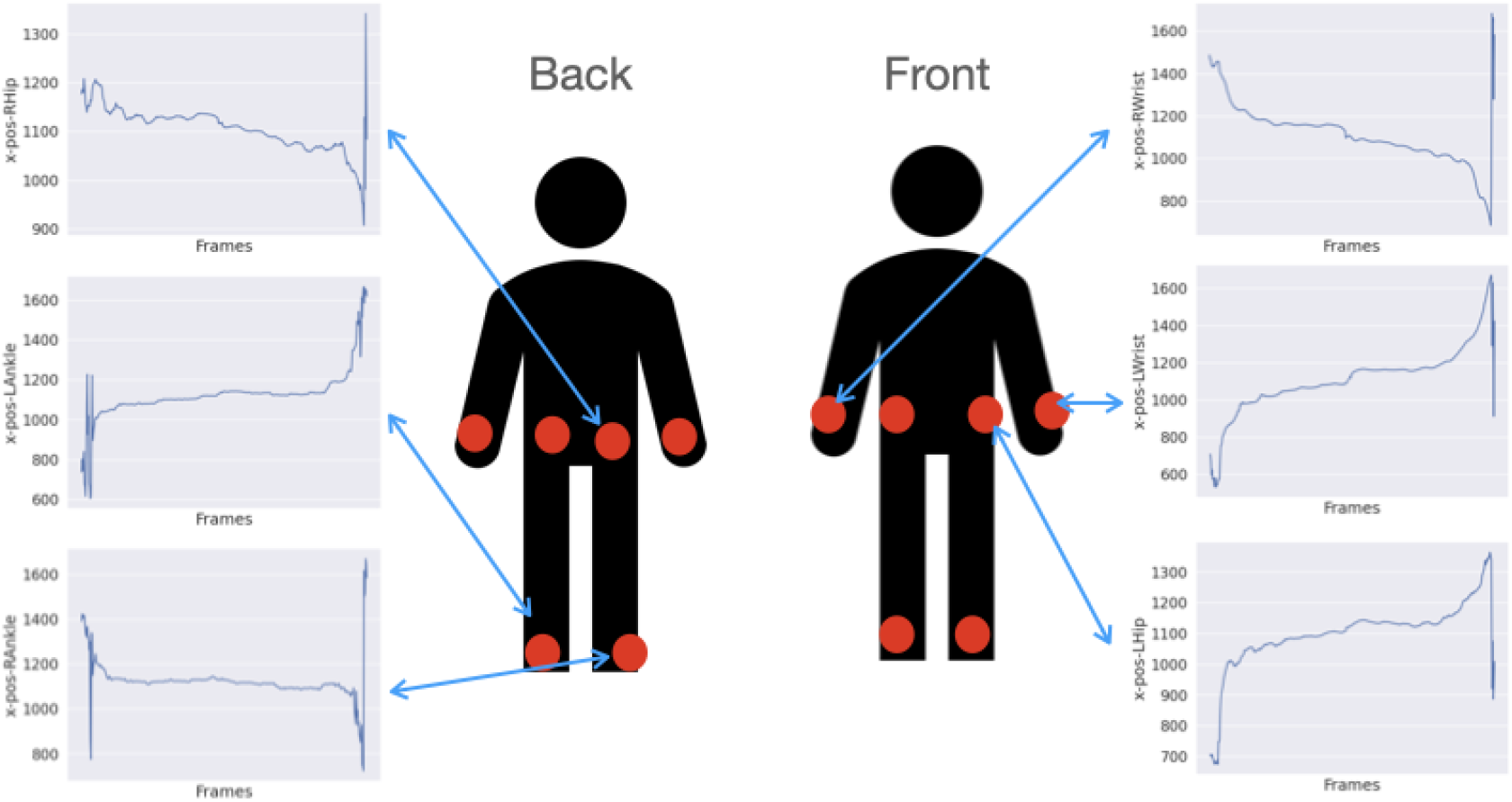
*X-pos* markers. Illustration of the body markers used to create the time series *X-pos*.

**Figure S7:**
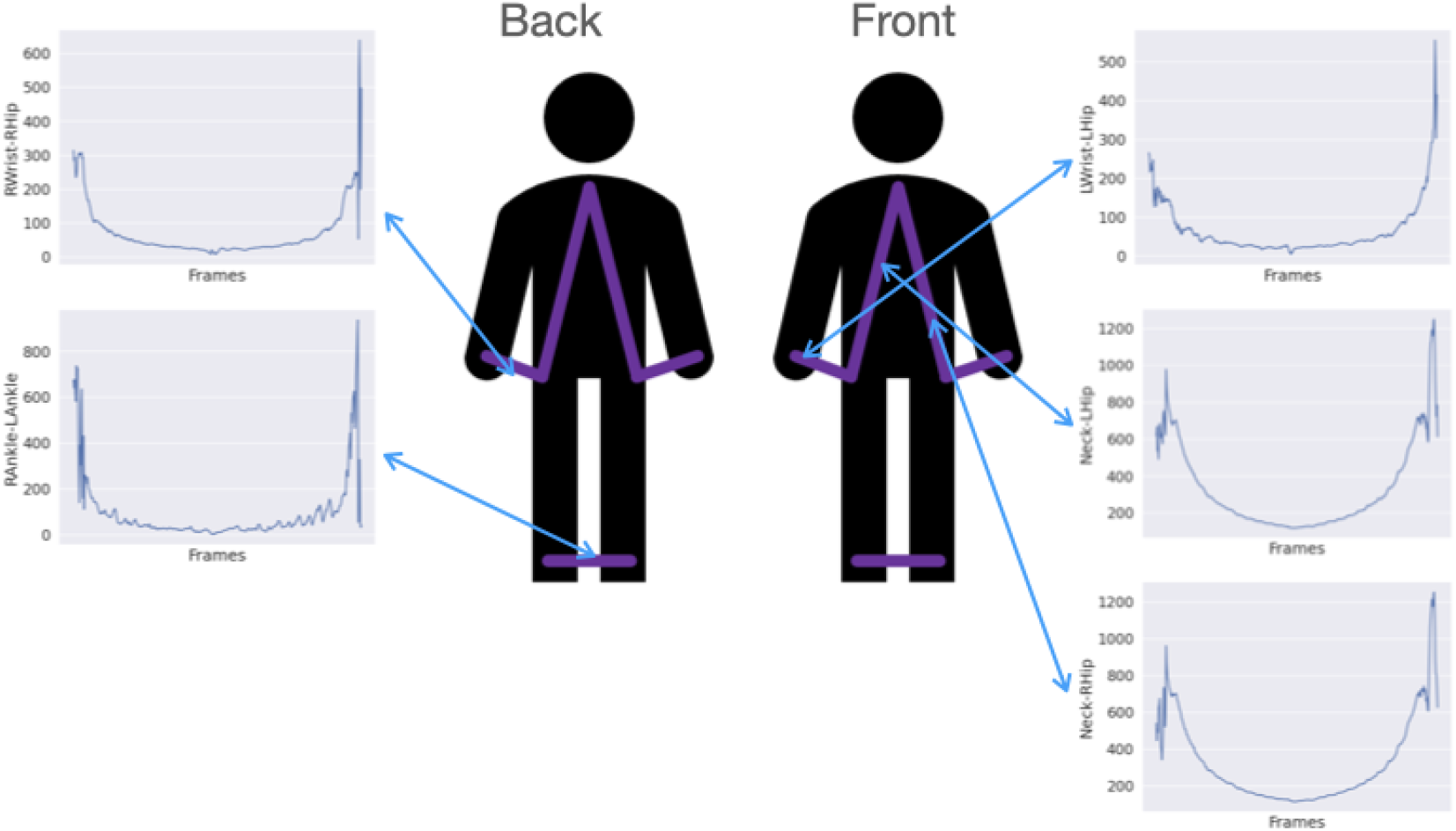
*Dist* markers. Illustration of the distances between body markers used to create the time series *Dist*.

**Figure S8:**
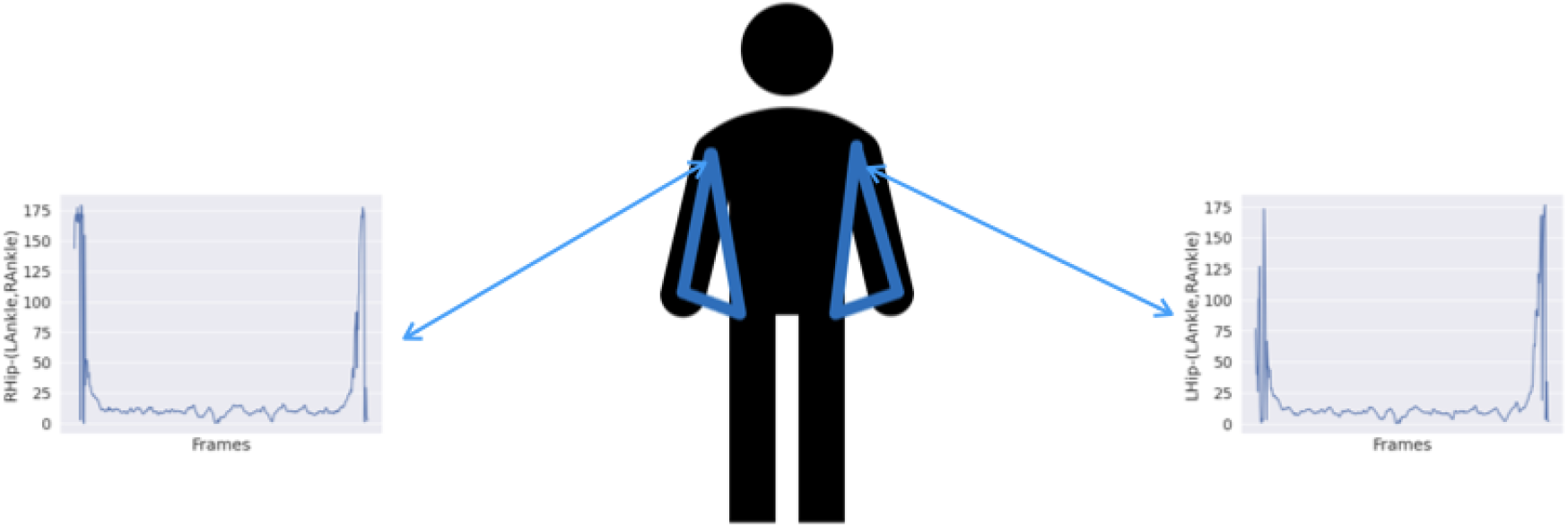
*Upper* markers. Illustration of the angles in the upper body used to create the time series *Upper*.

**Figure S9:**
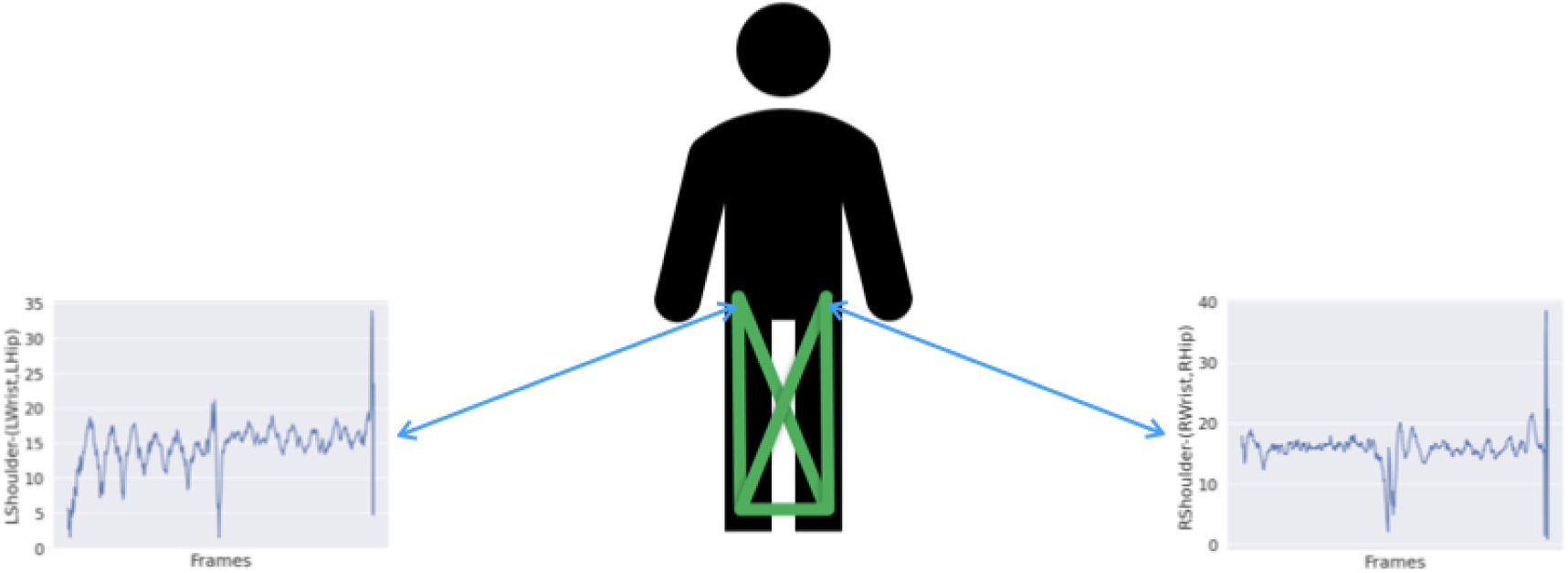
*Lower* markers. Illustration of the angles in the lower body used to create the time series *Lower*.

